# ChatGPT-o1 and the Pitfalls of Familiar Reasoning in Medical Ethics

**DOI:** 10.1101/2024.09.25.24314342

**Authors:** Shelly Soffer, Vera Sorin, Girish N Nadkarni, Eyal Klang

## Abstract

Large language models (LLMs) like ChatGPT often exhibit Type 1 thinking—fast, intuitive reasoning that relies on familiar patterns—which can be dangerously simplistic in complex medical or ethical scenarios requiring more deliberate analysis. In our recent explorations, we observed that LLMs frequently default to well-known answers, failing to recognize nuances or twists in presented situations. For instance, when faced with modified versions of the classic “Surgeon’s Dilemma” or medical ethics cases where typical dilemmas were resolved, LLMs still reverted to standard responses, overlooking critical details. Even models designed for enhanced analytical reasoning, such as ChatGPT-o1, did not consistently overcome these limitations. This suggests that despite advancements toward fostering Type 2 thinking, LLMs remain heavily influenced by familiar patterns ingrained during training. As LLMs are increasingly integrated into clinical practice, it is crucial to acknowledge and address these shortcomings to ensure reliable and contextually appropriate AI assistance in medical decision-making.

## Introduction

Large language models (LLMs) have revolutionized natural language processing by enabling machines to generate human-like text. Despite their advanced capabilities, LLMs often exhibit Type 1 thinking—fast, intuitive reasoning that relies heavily on familiar patterns and past experiences ^1,2^. While this can be efficient, it poses risks in medical and ethical contexts that require careful analysis and consideration of nuanced information, known as Type 2 thinking.

## Methods

We conducted a series of tests on several LLMs, including ChatGPT-o1, which is designed for enhanced analytical reasoning. The models were presented with a set of lateral thinking puzzles and medical ethics scenarios that included intentional twists to challenge default assumptions. For instance, in the modified “Surgeon’s Dilemma,” explicit details were provided to invalidate the typical solution. In medical ethics cases, scenarios were constructed where usual dilemmas were already resolved within the prompt.

## Results

The LLMs frequently defaulted to familiar solutions, overlooking critical details that required a different response. As shown in **Table 1**, in the modified “Surgeon’s Dilemma,” despite information that the father was the surgeon and the mother was a social worker, and only the boy was in the accident, the models still concluded that the surgeon was the mother. In medical ethics scenarios presented in **Table 2**, even when patients had already disclosed critical information or parental consent was granted, the models continued to discuss standard ethical debates as if these issues were unresolved. Notably, ChatGPT-o1 showed limited improvement over its predecessors, successfully recognizing twists in only a few instances.

## Discussion

In these explorations with LLMs, we demonstrateed a recurring pattern: LLMs fail to recognize nuances or twists, defaulting instead to familiar answers that are not appropriate for the situation at hand.

Even when trained for Type 2 like thinking (“Chain of Thoughts”), LLMs may default to sequences they have identified as highly probable during training, especially frequently recurring statements such as clichéd puzzles or certain very familiar ethical dilemmas^3^. As a result, they may apply familiar patterns even when these are not entirely suitable for the specific context of the prompt^4^.

While human Type 1 thinking is efficient and often reliable, it is also adaptive and shaped by emotional and contextual understanding. Humans may recognize when a situation requires more deliberate, analytical thought—Type 2 thinking—and can shift their approach accordingly. Similarly, OpenAI recently introduced ChatGPT-o1 model, designed to spend more time thinking before answering, aiming to reason through complex tasks and solve harder problems than previous models. While this represents a step toward Type 2 thinking, our observations indicate that these models still require further refinement to handle nuanced scenarios effectively.

As LLMs are being integrated into medical practice^5,6^, it is important to recognize these limitations despite their intended enhancements. The risk of pattern reliance, where the model’s outputs are overly influenced by heavily repeated training examples (over-fitting), underscores the need for critical evaluation of AI-generated responses.

## Conclusion

Even with advancements toward Type 2 thinking, heavily repeated training data can influence LLMs’ clinical decisions. It is important to recognize this behavior before integrating the technology into clinical use.

## Data Availability

All data produced in the present work are contained in the manuscript

