## Supplementary material for "ChatGPT-o1 and the Pitfalls of Familiar Reasoning in Medical Ethics": Table 1

Table 1: Examples of Lateral Thinking Puzzles Where LLMs Failed to Recognize Twists

|  | **Original Question** | **Original Answer** | **Rephrased Question** | **GPT4o Answer** | **GPTo1 answer** | **Llama-3.1-70b Answer** | **Claude Sonnet 3.5** |
| --- | --- | --- | --- | --- | --- | --- | --- |
| **1** | **The Surgeon's Dilemma**: A father and his son are in a horrible car crash that kills the dad. The son is rushed to the hospital. Just as he's about to go under the knife, the surgeon says, "I can't operate—that boy is my son!" How can this be? | The surgeon is the boy’s mother. | A son has a father who is a surgeon and a **mother who is a social worker**. The son is rushed to the hospital. Just as he's about to go under the knife, the surgeon says, "I can't operate—that boy is my son!" How can this be? | The surgeon who says, "I can't operate—that boy is my son!" is the boy's mother.  In this scenario, the mother is both a social worker and a surgeon. This addresses the implicit bias that might assume the surgeon must be male. | The surgeon is the boy's mother. Despite the initial description stating that his mother is a social worker, it's possible for her to hold both professions or for the term "mother" to refer to another maternal figure. This riddle challenges the assumption that a surgeon must be male and highlights potential unconscious biases. | This is a classic lateral thinking puzzle!  The answer is that the surgeon is the boy's mother.  The puzzle's wording is carefully crafted to make us assume the surgeon is a man, but it doesn't explicitly state that. Since the boy's mother is a social worker, it's entirely possible that the surgeon is the boy's mother, making it a perfectly plausible and heartwarming twist! | The surgeon is the boy's mother.  This riddle plays on gender stereotypes and assumptions about professions. Many people might initially assume the surgeon is male, but in this case, the surgeon is actually the boy's mother.  Would you like me to explain the riddle further or discuss why this type of puzzle can be effective at revealing unconscious biases? |
| **2** | **The Window Cleaner**: A window cleaner is cleaning the windows on the 25th floor of a skyscraper when he slips and falls. He is not wearing a safety harness and nothing slows his fall, yet he is not hurt. How can this be? | He was cleaning the inside of the windows. | A window cleaner is cleaning the windows on the 25th floor of a skyscraper when he slips and falls. He is not wearing a safety harness and nothing slows his fall, **and he dies**. How can this be? | The window cleaner slips and falls inside the building. He is cleaning the windows from the inside, not the outside. Thus, when he slips and falls, he falls inside the room. | The window cleaner was cleaning the windows **inside** the building on the 25th floor. While working, he slipped and fell **down an open staircase or elevator shaft within the building**. Since he was inside, he wasn't wearing a safety harness, and nothing slowed his fall. Unfortunately, the fall was fatal. | A classic lateral thinking puzzle!  The answer is that the window cleaner was inside the building, cleaning the windows from the inside, when he slipped and fell. Since he was already inside, he didn't have far to fall, and his fall was likely onto the floor of the 25th story, which would be fatal without additional injuries.  No need for a safety harness when you're cleaning from the inside! | Let's think through this step-by-step:  1. The scenario describes a window cleaner falling from the 25th floor of a skyscraper.  2. We're told that he's not wearing a safety harness and nothing slows his fall.  3. The question states that he dies, which seems expected given the height of the fall.  4. However, the question "How can this be?" suggests there's a twist or unexpected element we need to consider.  Given these facts, the most likely explanation is:  The window cleaner was cleaning the windows from the inside of the building, not the outside. When he slipped and fell, he fell onto the floor of the 25th story, not off the building.  This explanation accounts for all the given information:  - He was indeed on the 25th floor.  - He wasn't wearing a safety harness (which wouldn't be necessary inside).  - Nothing slowed his fall (as he fell directly onto the floor).  - He died from the fall, which could happen even from a standing height if he hit his head or landed in an unfortunate way.  The key to this riddle is the assumption most people make that window cleaners always work from the outside of the building, which isn't necessarily the case, especially for very tall structures. |
| **3** | **The Fox, Chicken, and Corn Puzzle:** A farmer needs to transport a fox, a chicken, and a sack of corn across a river. He has a boat but can only take one at a time. He cannot leave the fox alone with the chicken, or the chicken with the corn. How does he do it? | The farmer takes the chicken across first. He returns alone and takes the fox across next. He brings the chicken back with him, leaves the chicken, and takes the corn across. Finally, he returns alone to fetch the chicken. | A farmer needs to transport a fox, a chicken, and a sack of corn across a river. He has a boat but can only take **two** at a time. He cannot leave the fox alone with the chicken, or the chicken with the corn. How does he do it? | To solve this puzzle, the farmer needs to make several trips across the river, ensuring at no point are the chicken and the corn or the fox and the chicken left alone together on one side of the river. Here's a step-by-step solution:  First Trip: The farmer takes the chicken across the river and leaves it on the other side.  Second Trip: The farmer returns alone to the original side and takes the fox across the river.  Third Trip: The farmer leaves the fox on the other side, but takes the chicken back with him to the original side.  Fourth Trip: The farmer leaves the chicken and takes the corn across the river to the other side.  Fifth Trip: Finally, the farmer returns alone to the original side and takes the chicken across the river.  Now, all three items — the fox, the chicken, and the sack of corn — are safely on the other side of the river, and none were left alone with another that could cause a problem. This sequence prevents any issues of the chicken being with the corn or the fox with the chicken without supervision. | To solve this classic river crossing puzzle, the farmer must carefully plan his trips to ensure that the fox, chicken, and corn are never left alone in incompatible pairs. Here’s a step-by-step guide:   1. **First Trip**: The farmer takes the **chicken** across the river.    - **Left Bank**: Fox, Corn    - **Right Bank**: Chicken 2. **Second Trip**: The farmer returns alone to the left bank.    - **Left Bank**: Fox, Corn, Farmer    - **Right Bank**: Chicken 3. **Third Trip**: The farmer takes the **fox** across the river.    - **Left Bank**: Corn    - **Right Bank**: Fox, Chicken 4. **Fourth Trip**: The farmer brings the **chicken** back to the left bank.    - **Left Bank**: Chicken, Corn    - **Right Bank**: Fox 5. **Fifth Trip**: The farmer takes the **corn** across the river.    - **Left Bank**: Chicken    - **Right Bank**: Fox, Corn 6. **Sixth Trip**: The farmer returns alone to the left bank.    - **Left Bank**: Chicken, Farmer    - **Right Bank**: Fox, Corn 7. **Seventh Trip**: The farmer takes the **chicken** across the river again.    - **Left Bank**: None    - **Right Bank**: Fox, Chicken, Corn   Throughout this process, the farmer ensures that:   - The **fox and chicken** are never left alone together without the farmer. - The **chicken and corn** are never left alone together without the farmer.   By following these steps, all three items are safely transported across the river. | This is a classic puzzle! Here's the solution:  **Step 1:** The farmer takes the chicken and the corn across the river first, leaving the fox behind.  **Step 2:** The farmer goes back to the initial side and picks up the fox.  **Step 3:** The farmer takes the fox across the river and leaves it on the other side with the corn.  **Step 4:** The farmer goes back to the initial side and picks up the chicken (which was left there in step 2).  **Step 5:** The farmer takes the chicken across the river again and reunites it with the fox and the corn.  This solution ensures that the fox and the chicken are never left together, and neither are the chicken and the corn. | Let's approach this problem step by step:   1. Initial state: All items (farmer, fox, chicken, corn) are on one side of the river. 2. First trip:    - Farmer takes the chicken across (fox and corn are safe together)    - Farmer returns alone 3. Second trip:    - Farmer takes the fox across    - Farmer brings the chicken back (can't leave fox and chicken together) 4. Third trip:    - Farmer takes the corn across (chicken is alone, which is safe)    - Farmer returns alone 5. Fourth trip:    - Farmer takes the chicken across 6. Final state: All items are safely on the other side of the river.   This solution ensures that at no point is the fox left alone with the chicken, or the chicken with the corn.  Would you like me to explain any part of this solution in more detail? |
| **4** | **The Two Guards Puzzle:** You reach a fork in the road guarded by two guards. One path leads to safety, the other to certain death. One guard always tells the truth, and the other always lies, but you don't know which is which. You can ask one question to one guard to find out which path leads to safety. What do you ask? | Ask either guard, "If I asked the other guard which path leads to safety, what would he say?" Take the opposite path of whatever answer you receive. This works because whether you ask the liar or the truth-teller, they will both point you to the dangerous path, so you should choose the opposite. | You reach a fork in the road guarded by two guards. One path leads to safety, the other to certain death. One guard always tells the truth, and the other **always tells the truth**, but you don't know which is which. You can ask one question to one guard to find out which path leads to safety. What do you ask? | You ask one guard the following question:  "If I were to ask the other guard which path leads to safety, what would he say?"  Then, take the opposite path of the one mentioned in the guard's answer.  Here’s the reasoning:  If you ask the truth-telling guard, he will truthfully tell you what the lying guard would say, which will be the incorrect path.  If you ask the lying guard, he will lie about what the truth-telling guard would say, also pointing to the incorrect path.  In both cases, the guard you ask will indicate the path that leads to certain death. Therefore, you should take the opposite path to reach safety. | Given the information that **both guards always tell the truth**, you can simply ask either guard which path leads to safety. Since they both always tell the truth, their answer will be accurate.  **Question to ask:** "Which path leads to safety?" | Classic puzzle!  You can ask one of the guards: "If I were to ask the other guard which path leads to safety, what would they say?"  Think about it for a moment...  If you ask the truth-telling guard, they will tell you that the lying guard would point to the dangerous path.  If you ask the lying guard, they will lie about what the truth-telling guard would say, and point to the dangerous path.  So, regardless of the answer, you can safely assume the opposite path leads to safety! | This is a classic logic puzzle! Let's think through it step by step to find the optimal question to ask.   1. First, we need to recognize that asking a direct question like "Which path leads to safety?" won't work, because we don't know if we're asking the truthful guard or the lying guard. 2. The key is to formulate a question that will give us the correct answer regardless of which guard we ask. 3. One effective strategy is to ask a question that involves what the other guard would say. This way, we're incorporating both guards' responses into a single question. 4. The optimal question to ask is:   "If I were to ask the other guard which path leads to safety, what would he say?"   5. Let's break down why this works:    - If you ask the truthful guard, he will truthfully tell you what the lying guard would say, which would be the wrong path.    - If you ask the lying guard, he will lie about what the truthful guard would say, which would also result in the wrong path. 6. Therefore, whichever answer you get, you should choose the opposite path.   By using this question, you're guaranteed to identify the safe path, regardless of which guard you ask. This puzzle demonstrates the power of logical reasoning and how carefully constructed questions can extract truth even in situations with limited and potentially misleading information. |
| **5** | **Two Fathers and Two Sons Go Fishing**: Two fathers and two sons go fishing. Each of them catches one fish. So why do they bring home only three fish? | There are only three people: a grandfather, his son, and his grandson. | Two fathers and two sons go fishing. Each of them catches one fish. So why do they bring home **four fish**? | This riddle can be explained by the relationships among the fishermen. Let's break it down:  There are two fathers and two sons, but they are not four distinct people.  Instead, they can be:  A grandfather (who is a father),  His son (who is both a son and a father),  And the son's son (who is a son).  In this way, we have:  The grandfather: 1 father.  The father: 1 son (to the grandfather) and 1 father (to the grandson).  The grandson: 1 son.  Counting them individually:  Grandfather (1 person)  Father (1 person)  Son (1 person)  Therefore, there are only three people, but they still meet the criteria of being two fathers and two sons. Since each catches one fish, they bring home a total of three fish, not four. The riddle's tricky wording can lead to confusion, but it's clear with this explanation. | They bring home four fish because there are four people: two fathers and two sons. This situation involves two separate father-son pairs, making up four individuals in total. Each person catches one fish, resulting in four fish being brought home.  **Explanation:**   - **Person 1:** Father A (father of Son A) - **Person 2:** Son A (son of Father A) - **Person 3:** Father B (father of Son B) - **Person 4:** Son B (son of Father B)   This accounts for two fathers and two sons, and each of the four individuals catches one fish, totaling four fish.  **So, they bring home four fish because there are four people—each a father or a son—who each caught one fish.**  Because there are four people—two fathers and two sons—so each catches a fish, totaling four fish. | Clever puzzle!  The answer is that one of the fathers is also a son. In other words, there are three generations of a family: a grandfather, a father, and a son. So, two fathers (the grandfather and the father) and two sons (the father and the son) go fishing, and each of them catches one fish, making a total of three fish. But wait, you asked why they bring home four fish? The answer is that the puzzle statement is incorrect, they actually bring home three fish! | Let's think through this step-by-step:   1. The problem states that there are two fathers and two sons, which might initially make us think there are four people. 2. However, the key is to realize that these relationships can overlap. 3. Consider a three-generation scenario:    - A grandfather (who is a father)    - A father (who is both a son and a father)    - A son 4. In this case:    - The grandfather is a father    - The father is both a son (to the grandfather) and a father (to the son)    - The son is a son 5. So we actually have only three people, but they fulfill the roles of "two fathers and two sons":    - Two fathers: grandfather and father    - Two sons: father and son 6. Each of these three people catches one fish.   Therefore, they bring home three fish in total, not four. The puzzle's statement about bringing home four fish is a misdirection to make the problem more challenging. |
