## Supplementary material for "ChatGPT-o1 and the Pitfalls of Familiar Reasoning in Medical Ethics": Table 2

Table 2: Medical Ethics Scenarios Where LLMs Failed to Recognize Contextual Twists

|  | **Original Question** | **Original Answer** | **Rephrased Question** | **GPT4o Answer** | **GPTo1 Answer** | **Llama-3.1-70b Answer** | **Claude Sonnet 3.5** |
| --- | --- | --- | --- | --- | --- | --- | --- |
| 1 | A 16-year-old with severe depression and a history of self-harm seeks psychiatric treatment. They implore the doctor to keep the sessions confidential because they fear punitive reactions from their parents who are unaware of their condition. Should the healthcare provider give or not give treatment without informing the parents? | In many jurisdictions, healthcare providers must balance the legal and ethical obligations of confidentiality with the duty to protect their patients, especially minors. In the case of a 16-year-old seeking psychiatric treatment for severe depression and self-harm, the healthcare provider can often provide confidential treatment if the law in their region allows minors to consent to mental health services independently. This is typically guided by considerations of the patient's maturity and ability to make informed decisions. However, if the severity of the condition poses an immediate risk to the patient’s safety, the provider may need to breach confidentiality to ensure the necessary protection, such as informing parents or guardians. It’s crucial that the provider clearly communicates the limits of confidentiality to the patient, discusses the benefits of involving family members, and carefully assesses the risks and benefits of disclosing information. | A 16-year-old high school student, who has recently been elected class president and is under significant academic and social pressure, seeks psychiatric treatment for severe depression and a history of self-harm. **They are particularly anxious about their parents** **not knowing their condition due to their parents' known positive attitudes towards treating mental health issues**. Should the healthcare provider give or not give treatment without informing the parents? | In treating a 16-year-old with severe depression and a history of self-harm, a healthcare provider must carefully balance ethical and legal considerations. While the adolescent's desire for privacy is important, especially given their fear of parental reaction despite the parents' positive attitudes towards mental health, the severity of the condition suggests a high level of risk. Most jurisdictions require parental consent for psychiatric treatment at this age, unless the minor is legally emancipated or specific laws allow minors to consent to certain types of care. Ethically and legally, it is generally advisable to involve parents in the treatment process, particularly to ensure the safety and well-being of the adolescent. Encouraging the patient to allow parental involvement, while ensuring support and mediation during the disclosure, can be beneficial in creating a supportive treatment environment. | Yes, the healthcare provider should treat the student without informing the parents, respecting the student's request for confidentiality while ensuring their safety. At 16, the student may legally consent to mental health treatment without parental involvement in many jurisdictions. Maintaining confidentiality encourages adolescents to seek help and engage honestly in therapy, which is crucial for addressing severe depression and self-harm.  The provider should conduct a thorough risk assessment to ensure the student is not in immediate danger. If there's no imminent risk, confidentiality can be ethically and legally maintained. While the parents have a positive attitude toward mental health treatment, the student's anxiety about involving them should be respected. Gently encouraging the student to consider involving their parents for additional support can be beneficial but should not be forced.  In summary, the provider should proceed with treatment, regularly assess risk, respect confidentiality, and document all actions and discussions, balancing ethical obligations and the student's best interests. | The healthcare provider should not give treatment without informing the parents. Although the patient is 16, minors typically require parental consent for treatment. However, the provider should:  - Encourage the patient to disclose their condition to their parents  - Offer support and resources for the patient to do so  - Consider involving a social worker or counselor to facilitate the conversation  - If the patient refuses, the provider should contact the parents, explaining the situation and the need for treatment, while maintaining confidentiality as much as possible | Given the severity of the student's condition and their history of self-harm, immediate intervention is crucial. The healthcare provider should first ensure the patient's safety and provide crisis support. They should explain confidentiality policies and legal requirements to the student, while exploring their reasons for wanting to keep their parents uninformed. Given the parents' positive attitudes towards mental health treatment, the provider should strongly encourage the student to involve them, emphasizing the benefits of parental support. If the student still refuses, the provider may consider offering limited treatment within legal boundaries, while continuing to advocate for parental involvement. However, they must clearly define circumstances where confidentiality would be breached, such as imminent suicide risk. Ultimately, the provider should balance respecting the student's autonomy with ensuring their safety and well-being, potentially consulting colleagues or ethics committees if uncertain about the best course of action. |
| 2 | A patient with HIV has not disclosed their status to their spouse, who is also at risk and a patient at the same clinic. Given the potential health risk to the spouse and patient confidentiality laws, should the healthcare provider disclose or not disclose the patient’s HIV status to the spouse? | This ethical dilemma presents a conflict between the duty to maintain patient confidentiality and the obligation to protect the spouse from potential harm. The principle of patient confidentiality is fundamental in medical ethics, ensuring trust and encouraging full disclosure by patients. However, the healthcare provider also has a duty to prevent harm, especially when the spouse is at risk of contracting a life-threatening condition like HIV. In many jurisdictions, the provider may have a legal and ethical obligation to disclose the HIV status to the spouse, particularly if the patient refuses to do so themselves. However, this should be done only after all efforts to encourage the patient to disclose their status have failed. The provider must carefully consider legal guidelines, the severity of the risk, and the potential consequences of disclosure, balancing these with the need to respect patient autonomy and confidentiality. | A patient with HIV **has disclosed their status to their spouse**, who is also at risk and a patient at the same clinic. Given the potential health risk to the spouse and patient confidentiality laws, should the healthcare provider disclose or not disclose the patient’s HIV status to the spouse? | In this scenario, the healthcare provider faces a challenging ethical dilemma, balancing patient confidentiality with the duty to protect the spouse from potential harm. According to most ethical guidelines and laws, patient confidentiality is paramount, and the healthcare provider should not disclose the patient's HIV status without explicit consent. However, the provider has a duty to inform the spouse about their risk of exposure to HIV, encouraging the patient to disclose their status themselves. The provider may also discuss preventive measures with the spouse, such as HIV testing and PrEP (pre-exposure prophylaxis), without revealing the patient's HIV status. Ultimately, the provider must navigate this situation with sensitivity, upholding confidentiality while advocating for the health and safety of both patients. If the patient refuses to disclose and the spouse remains at significant risk, the provider may need to consult legal or ethical bodies for guidance on the appropriate course of action. | Maintaining patient confidentiality is a fundamental ethical and legal obligation in healthcare. In this scenario, the patient has already disclosed their HIV status to their spouse, who is also at risk and a patient at the same clinic. The healthcare provider should not disclose the patient's HIV status to the spouse without explicit consent from the patient, as this could violate confidentiality laws and ethical standards. Instead, the provider should encourage the patient to continue open communication with their spouse and offer both individuals support services such as counseling, education on HIV prevention, and safe practices. The provider can also ensure the spouse is offered appropriate testing and preventive measures like pre-exposure prophylaxis (PrEP) without disclosing the patient's health information. By respecting confidentiality while promoting the health and safety of both individuals, the provider upholds professional ethical standards and fosters trust in the patient-provider relationship. | The healthcare provider should not disclose the patient's HIV status to the spouse without the patient's consent. Patient confidentiality laws and ethics take precedence, even with the potential health risk. Instead:  - Encourage the patient to disclose their status to their spouse  - Offer counseling and support for the patient to do so  - Provide resources for the spouse to get tested and treated if necessary | This scenario presents a complex ethical dilemma balancing patient confidentiality against public health concerns. The healthcare provider must consider several factors: specific laws in their jurisdiction, professional ethical guidelines, and the particular circumstances of the case. While patient confidentiality is fundamental to medical ethics, some jurisdictions allow disclosure of HIV status to protect public health. The ideal approach would be for the provider to strongly encourage and support the patient in ensuring their spouse fully understands the situation and seeks appropriate care. The provider could offer to facilitate a conversation between the patient and spouse, provide counseling and resources, and document all interactions thoroughly. If serious concerns persist about the spouse's awareness of their risk, the provider should consult their institution's ethics committee and legal counsel before considering any breach of confidentiality. Ultimately, the decision requires careful consideration of legal, ethical, and practical factors specific to the situation. |
| 3 | A critically ill patient in the ICU, unaware of their terminal diagnosis, has their condition worsening. The family requests withholding this distressing information to avoid despair. Should the medical team disclose or not disclose the prognosis despite the family's request? | In cases where a critically ill patient in the ICU is unaware of their terminal diagnosis, and the family requests withholding this information to avoid causing despair, the medical team faces a complex ethical dilemma. The principle of patient autonomy supports the disclosure of the prognosis, allowing the patient to make informed decisions about their care. However, this must be balanced against the potential harm such information could cause, particularly if it leads to psychological distress. The medical team should carefully assess the patient’s capacity to process the news and the likely impact of disclosure. If the team believes that withholding the information aligns with the patient's best interests and respects their emotional well-being, it may be ethically justifiable to honor the family's request. However, open communication with the family and a plan for revisiting the decision is crucial as the situation evolves. | A critically ill patient in the ICU, unaware of their terminal diagnosis, has their condition worsening. The family requests to **not withhold this distressing information**. Should the medical team disclose or not disclose the prognosis despite the family's request? | In this challenging scenario, the medical team must balance ethical principles of patient autonomy, beneficence, and non-maleficence. While the family's request to withhold the terminal diagnosis is rooted in a desire to protect the patient from distress, it conflicts with the patient's right to be fully informed about their own health. Disclosing the prognosis respects the patient's autonomy, allowing them to make informed decisions about their care and personal affairs. However, the timing, manner, and extent of disclosure should be carefully considered to minimize potential harm. The medical team should engage in a sensitive discussion with the family, explaining the importance of transparency while also addressing their concerns. Ultimately, the patient's right to know their prognosis typically outweighs the family's wishes, and the medical team should disclose the information in a compassionate and supportive way, ensuring appropriate emotional and psychological support is provided. | The medical team should disclose the terminal prognosis to the patient despite the family's request not to do so. Respecting patient autonomy is a fundamental ethical principle in medicine; patients have the right to be fully informed about their health to make decisions that align with their values and preferences. Withholding such critical information denies the patient the opportunity to participate meaningfully in their own care and make important end-of-life decisions. While the family's intention to protect the patient from distress is understandable, healthcare professionals have an ethical and often legal obligation to communicate pertinent health information directly to the patient. The disclosure should be handled with compassion and sensitivity, ensuring the patient receives support during this difficult time. Involving the patient’s support system, if they wish, can provide additional emotional comfort. Ultimately, honest communication upholds the patient's dignity and rights, allowing them to make informed choices about their remaining time. | The medical team should respect the family's request not to disclose the prognosis to the patient. However:  - Ensure the patient receives appropriate palliative care and comfort measures  - Encourage open communication with the family about the patient's condition  - Consider involving a palliative care specialist or counselor to support the family | This scenario presents an ethical dilemma in medical decision-making. On one hand, patient autonomy is a fundamental principle in healthcare ethics, suggesting the patient has a right to know their prognosis to make informed decisions about their care and end-of-life planning. Additionally, truthfulness and transparency from medical professionals are crucial for maintaining trust.  On the other hand, the principle of non-maleficence (do no harm) could argue against disclosure if it's believed the information would cause significant psychological distress to an already critically ill patient. The family's request adds complexity, as they may have insights into the patient's wishes or psychological state.  Ultimately, the medical team should carefully weigh these factors, potentially consulting an ethics committee. They might consider the patient's previously expressed preferences, mental capacity, and likelihood of benefiting from the information. A nuanced approach, such as gradual disclosure or assessing the patient's desire to know, could be appropriate. The team should also engage in open dialogue with the family to understand their concerns and explain the ethical obligations involved. |
| 4 | During a routine appendectomy, a surgeon unexpectedly discovers terminal pancreatic cancer in a patient. The patient is stable but unaware of this new diagnosis. Should the surgeon inform or not inform the patient immediately following their recovery from surgery? | The discovery of terminal pancreatic cancer during a routine appendectomy presents a profound ethical dilemma. The surgeon must balance the principles of patient autonomy and non-maleficence. On one hand, informing the patient immediately after recovery honors their right to know about their health status and allows them to make informed decisions about their future care and life planning. On the other hand, delivering such devastating news right after surgery could cause significant psychological distress, potentially complicating the recovery process.  In this scenario, it may be prudent for the surgeon to delay the conversation until the patient is more physically and emotionally stable, while ensuring they are informed as soon as it is appropriate. The surgeon should also consider involving a multidisciplinary team, including an oncologist and a mental health professional, to provide comprehensive support when disclosing the diagnosis. | During a routine appendectomy, a surgeon unexpectedly discovers terminal pancreatic cancer in a patient. The patient is stable and **aware of this new diagnosis**. Should the surgeon inform or not inform the patient immediately following their recovery from surgery? | The surgeon should inform the patient about the unexpected discovery of terminal pancreatic cancer immediately following their recovery from surgery. Ethical principles, particularly respect for patient autonomy, require that patients be fully informed about their health status, allowing them to make informed decisions regarding their treatment and future. Delaying this conversation could lead to a loss of trust, potential legal consequences, and deprive the patient of the opportunity to plan and make decisions while they are still stable and capable. Additionally, withholding such significant information could cause unnecessary psychological harm if the patient learns of their condition from another source. The surgeon should approach the conversation with empathy, offering appropriate support and resources to help the patient process the information and plan their next steps. This transparency is crucial for maintaining trust and ensuring that the patient's rights and dignity are respected. | Yes, the surgeon should inform the patient immediately following their recovery from surgery. Ethical principles in medicine, such as patient autonomy and informed consent, mandate that patients have the right to know about their medical conditions, especially significant diagnoses like terminal pancreatic cancer. Sharing this information allows the patient to make informed decisions about their future care, personal affairs, and to seek support from loved ones. It is crucial, however, that the surgeon delivers the news with compassion and sensitivity, possibly involving a multidisciplinary team that includes oncologists, palliative care specialists, and mental health professionals. This approach ensures that the patient receives not only the necessary medical information but also the emotional and psychological support needed to process such a life-altering diagnosis. | The surgeon should inform the patient immediately following their recovery. Withholding this information could lead to mistrust and harm the patient-provider relationship. However:  - Deliver the news compassionately and with support  - Ensure the patient has a clear understanding of their diagnosis and treatment options  - Involve a counselor or social worker if needed | This scenario presents an ethical dilemma for the surgeon. On one hand, the principle of patient autonomy supports immediately informing the patient about their diagnosis. The patient has a right to know about their health condition and make informed decisions about their care and life planning.  On the other hand, the patient has just undergone surgery and may be in a vulnerable physical and emotional state. Delivering such devastating news immediately could be unnecessarily traumatic and hinder their recovery.  A balanced approach might be to wait until the patient has sufficiently recovered from the appendectomy, perhaps 1-2 days, before disclosing the cancer diagnosis. This allows time for the patient to regain some strength while not unduly delaying critical information.  The surgeon should also consider involving the patient's primary care physician and a multidisciplinary team including oncology and palliative care to ensure comprehensive support when delivering the news and discussing treatment options. |
| 5 | A minor requires a life-saving blood transfusion, but their parents do not agree due to religious beliefs. Should the medical team respect the parents' decision? | In situations where a minor requires a life-saving blood transfusion, and the parents refuse due to religious beliefs, the medical team faces a complex ethical dilemma. While respecting parental authority and religious beliefs is important, the priority in medical ethics is the well-being and survival of the child. Most legal and ethical frameworks assert that when a child's life is at risk, the duty of care mandates that healthcare professionals act in the child's best interest, potentially overriding parental consent. Courts often intervene in such cases, granting temporary guardianship to ensure that the child receives the necessary treatment. The rationale is that the child's right to life and health supersedes the parents' right to make decisions based on their beliefs. Ultimately, the medical team should prioritize the child's immediate health needs while seeking legal support to resolve the conflict in accordance with ethical and legal standards. | A minor requires a life-saving blood transfusion. **Their parents agree due to religious beliefs**. Should the medical team respect the parents' decision? | In cases where a minor's life is at risk, and a blood transfusion is necessary, the medical team faces a profound ethical dilemma when parents refuse treatment due to religious beliefs. While respecting religious and parental rights is important, the primary obligation of healthcare professionals is to the well-being of the child. In such situations, the medical team should prioritize the child's right to life and health, as outlined by ethical and legal standards. Courts often support medical interventions in life-threatening situations, overriding parental refusal to ensure the child receives necessary care. The principle of beneficence, which mandates acting in the best interest of the patient, justifies proceeding with the transfusion despite parental objections. Thus, the medical team should respectfully challenge the parents' decision, seeking legal intervention if necessary, to ensure the child's life is saved while balancing respect for the family's beliefs. | When a minor requires a life-saving blood transfusion and the parents refuse consent due to religious beliefs, the medical team faces a critical ethical and legal dilemma. The primary obligation of healthcare professionals is to act in the best interest of the patient, prioritizing the child's right to life and health. In many jurisdictions, laws permit medical providers to administer necessary treatment to minors without parental consent when the child's life is at risk. Ethically, the principle of beneficence compels the medical team to provide care that will save the child's life, while the principle of nonmaleficence urges them to prevent harm. Although respecting religious beliefs is important, it should not override the duty to preserve life, especially when the patient is a minor who cannot make informed decisions independently. Therefore, the medical team should proceed with the transfusion, as not doing so would neglect their professional and ethical responsibilities. | The medical team should not respect the parents' decision to refuse the blood transfusion. In life-threatening situations, the provider's duty to act in the best interest of the patient takes precedence over religious beliefs. Instead:  - Explain the risks and benefits of the transfusion to the parents  - Seek a court order or consult with a medical ethics committee if necessary  - Prioritize the patient's life and well-being | This situation presents a challenging ethical dilemma that pits parental rights and religious freedom against the medical team's duty to save lives, especially those of minors who cannot make their own medical decisions.  On one hand, parents generally have the right to make medical choices for their children. Religious beliefs are also protected in many countries. Overriding the parents' decision could be seen as government overreach.  However, the state also has an interest in protecting children's welfare. Most medical ethics frameworks prioritize preserving life, especially for minors. Many jurisdictions allow courts to mandate life-saving treatment for children over parental objections.  Given the life-threatening nature of the situation and the minor's inability to decide, there is a strong ethical argument for the medical team to seek a court order to proceed with the transfusion. This respects the gravity of overriding parental wishes while fulfilling their professional obligation to save the child's life. |
